# Predicting Long COVID in the National COVID Cohort Collaborative Using Super Learner

**DOI:** 10.1101/2023.07.27.23293272

**Authors:** Zachary Butzin-Dozier, Yunwen Ji, Haodong Li, Jeremy Coyle, Junming (Seraphina) Shi, Rachael V. Philips, Andrew Mertens, Romain Pirracchio, Mark J. van der Laan, Rena Patel, John M. Colford, Alan E. Hubbard, the National COVID Cohort Collaborative (N3C) Consortium

## Abstract

Post-acute Sequelae of COVID-19 (PASC), also known as Long COVID, is a broad grouping of a range of long-term symptoms following acute COVID-19 infection. An understanding of characteristics that are predictive of future PASC is valuable, as this can inform the identification of high-risk individuals and future preventative efforts. However, current knowledge regarding PASC risk factors is limited. Using a sample of 55,257 participants from the National COVID Cohort Collaborative, as part of the NIH Long COVID Computational Challenge, we sought to predict individual risk of PASC diagnosis from a curated set of clinically informed covariates. We predicted individual PASC status, given covariate information, using Super Learner (an ensemble machine learning algorithm also known as stacking) to learn the optimal, AUC-maximizing combination of gradient boosting and random forest algorithms. We were able to predict individual PASC diagnoses accurately (AUC 0.947). Temporally, we found that baseline characteristics were most predictive of future PASC diagnosis, compared with characteristics immediately before, during, or after COVID-19 infection. This finding supports the hypothesis that clinicians may be able to accurately assess the risk of PASC in patients prior to acute COVID diagnosis, which could improve early interventions and preventive care. We found that medical utilization, demographics, anthropometry, and respiratory factors were most predictive of PASC diagnosis. This highlights the importance of respiratory characteristics in PASC risk assessment. The methods outlined here provide an open-source, applied example of using Super Learner to predict PASC status using electronic health record data, which can be replicated across a variety of settings.

## BACKGROUND

As the mortality rate associated with acute COVID-19 incidence wanes, investigators have shifted focus to determining its longer-term, chronic impacts (1). Post-acute Sequelae of COVID-19 (PASC) is a loosely categorized consequence of acute infection that is related to dysfunction across multiple biological systems (2). Much remains unknown about PASC, leaving individuals uncertain regarding their risk for PASC and what factors may contribute to this risk. Prediction of individual risk for PASC diagnosis can allow us to identify what populations are at the greatest risk for PASC, and interpretation of these predictors may generate hypotheses regarding underlying drivers of PASC incidence.

Electronic health record (EHR) databases, such as the National COVID Cohorts Collaborative (N3C), provide an important tool for predicting, evaluating, and understanding PASC (3,4). There is a broad range of PASC symptoms, diagnostic criteria, and hypothesized causal mechanisms, which has made it difficult for investigators to build generalizable predictions (5–7). Given this heterogeneity, multi-site evaluations including large sample sizes and high-dimensional covariate information can provide opportunities to build models that can accurately predict PASC risk.

Due to the broad range of factors associated with PASC, the high dimensionality of the large EHR databases, and the unknown determinants of Long COVID, modeling methods for predicting PASC must be highly flexible. Super Learner (SL) is a flexible, ensemble (stacked) machine learning algorithm that uses cross-validation to learn the optimal weighted combination of a specified set of algorithms (8,9). The SL is grounded in statistical optimality theory that guarantees it will perform at least as well as the best-performing algorithm included in the library for large sample sizes. Thus, a rich library of learners, with a sufficient sample size, will ensure optimal performance. The SL can be specified to maximize any performance metric, such as mean squared error (9). Given the large sample size of high-dimensional data in EHR databases, SL is well positioned to predict individual risk of PASC diagnosis in this setting.

Here, we used the SL to predict PASC diagnosis in COVID-infected patients, given a diverse set of features curated from the EHR. We also investigated the importance of features for predicting PASC by assessing the importance of each individual feature, and by assessing groups of features based on temporality (baseline, pre-COVID, acute COVID, and post-COVID features), and by hypothesized clinical domains of PASC.

## METHODS

### Sample

The Long COVID Computational Challenge (L3C, DUR RP-5A73BA) sample population was selected from the N3C dataset, a national, open dataset that has been described previously (3,4). N3C has created a centralized repository where investigators can access and analyze data from more than 7 million COVID-19 patients, including 27 billion rows of data, from 80 sites across the United States while maintaining patient privacy (10,11). The L3C sample included cases of patients diagnosed with PASC (ICD code U09.9) and matched controls with a documented COVID-19 diagnosis who had at least one medical visit more than 4 weeks after their initial COVID diagnosis date. ICD Code U09.9, which was established on October 1, 2022, indicates a diagnosis for reimbursement purposes and enables linkage with COVID-19 diagnosis for patients experiencing post-acute sequelae of infection (12). Controls were selected at a 1:4 (case:control) ratio and were matched based on the distribution of medical visits prior to COVID-19 diagnosis. The primary outcome of interest was PASC diagnosis via ICD code U09.9. In order to evaluate our model’s predictive accuracy, we used a 10% holdout test set

### Feature selection

Our set of features for predicting PASC included those previously described in the literature (3) and additional features related to subject-matter knowledge and patterns of missingness. We extracted 304 features from N3C data. After indexing across four time periods (more below) and transforming features into formats amenable to machine learning analysis, our sample included 1,339 features (see Supplemental Table 1. Metadata). Details regarding feature selection and processing can be accessed via GitHub (https://github.com/BerkeleyBiostats/l3c_ctml/tree/v1). For continuous features, we included the minimum, maximum, and mean values for each measurement in each temporal window. For binary features, we either included an indicator (when repetition was not relevant) or a count (when repetition was relevant) over each time period and we re-coded categorical variables as indicators.

#### Temporal windows

We divided each participant’s records into four temporal windows: baseline, which consisted of all records occurring a minimum of 37 days before the COVID index date (*t* - 37, where *t* represents the COVID index date), and all time-invariant factors (such as sex, ethnicity, etc.); pre-COVID, observations falling between 37 and 7 days prior to the index date (*t* - 37 to *t* - 7); acute COVID, observations falling 7 days prior to 14 days after to the index date (*t* - 7 to *t* + 14); and post-COVID, records from 14 to 28 days after the index date (*t* + 14 to *t* + 28). The acute COVID window begins 7 days prior to the reported infection date, in order to conservatively include early COVID symptoms prior to official diagnosis.

#### Features described in the literature

We extracted and transformed key features that were identified in prior research using N3C data as risk factors for PASC.(3) These features included 199 previously described factors related to medical history, diagnoses, demographics, and comorbidities (3).

#### Temporality

To account for differences in follow-up, we included as an additional factor a continuous variable for follow-up time, defined as the number of days between the COVID index date and the most recent observation. To account for temporal trends of COVID (such as seasonality and dominant variant), we included categorical (ordinal) covariates for the season and months since the first observed COVID index date.

#### Missing data

To avoid dropping any observations, we imputed missing observations with 0 values and added indicator variables for imputed values.(13) By using flexible ensemble machine learning, which allows for interactions between imputed variables and the missingness indicators, we allow the patterns of missingness to be potential predictors of PASC.

#### COVID-19 positivity

We added several measures of COVID severity and persistent SARS-CoV-2 viral load, which are associated with PASC incidence (14). We imported measures of COVID-19 severity as well as 15 measures of COVID-19 infection from laboratory measurements, which provided insights into persistent SARS-CoV-2 viral load. We assessed the duration of COVID-19 viral positivity separately for each laboratory measure of COVID-19 and each temporal window. For participants who had both a positive and negative value of a given test during a temporal window, we took the midpoint between the last positive test and the first negative test as being the endpoint of their positivity. For individuals who had a positive test but no subsequent negative test within that temporal window, we determined their endpoint to be their final positive test plus three days. We included separate missingness indicators in each temporal window for each test, for a positive value for each test, and for a negative value following a positive value to indicate an imputed positivity endpoint. We included the calendar date of index infection to account for the COVID-19 viral strain, given our lack of variant data.

#### Additional features

We incorporated the laboratory measurements related to anthropometry, nutrition, COVID positivity, inflammation, tissue damage due to viral infection, auto-antibodies and immunity, cardiovascular health, and microvascular disease, which are potential predictors of PASC (14). We also extracted information about smoking status, alcohol use, marital status, and use of insulin or anticoagulant from the observation table as baseline characteristics of individuals. We included the number of times a person has been exposed to respiratory devices (e.g. supplemental oxygen, ventilator) in each of the four windows from the device table. We extracted covariates related to COVID severity, vaccination history, demographics, medical history, and previous diagnoses from before and during acute COVID-19 infection.

### Prediction using ensemble machine learning

We used the SL, an ensemble machine learning method, also known as stacking, to learn the optimally weighted combination of candidate algorithms for maximizing the AUC. We reprogrammed the SL in Python in order to capitalize on the resources available in the N3C Data Enclave (e.g., PySpark parallelization), and this software is available to external researchers (https://github.com/BerkeleyBiostats/l3c_ctml/tree/v1). We used an ensemble of four learners (a mix of parametric models and machine learning models): 1. Logistic regression; 2. L1 penalized logistic regression (with penalty parameter lambda = 0.01); 3. Gradient boosting (with n_estimators = 200, max_depth = 5, learning_rate = 0.1); 4. Random forest (max_depth = 5, num_trees = 20). The original candidate learner library consisted of a large set of candidate learners with different combinations of hyperparameters (e.g. gradient boosting (with n_estimators = [200, 150, 100, 50], max_depth = [3, 5, 7], learning_rate = [0.05, 0.1, 0.2]).

One important decision for optimizing an algorithm is to choose the metric that will be used to evaluate the fit and optimize the weighting of the algorithms in the ensemble. We used an approach developed specifically for maximizing the area under the curve (AUC) (15). The SL was specified such that it learned the combination of algorithms, including variations of gradient boosting and random forest, that maximized the AUC (15). Specifically, we used an AUC maximizing meta-learner with Powell optimization to learn the convex combination of these four candidate algorithms (15). The SL was implemented with a V-fold/k-fold cross-validation scheme with 10 folds.

### Prediction Performance

One important decision for optimizing an algorithm is to choose the metric that will be used to evaluate the fit and optimize the weighting of the algorithms in the ensemble. We used an approach developed specifically for maximizing the area under the curve (AUC) (15). Specifically, we used an AUC maximizing meta-learner with Powell optimization to learn the convex combination of these four candidate algorithms (15). The SL was implemented with a V-fold/k-fold cross-validation scheme with 10 folds.

### Variable importance

For the sake of computational efficiency, we worked with the discrete SL selector (the single candidate learner in the library with the highest cross-validated AUC) instead of the entire ensemble SL. In this case, the gradient-boosting learner was the candidate learner with the highest cross-validated AUC. We used a general approach (for any machine learning algorithm) known as Shapley values (16). We generated these values within three groupings of predictors for ease of interpretability: individual features (e.g. cough diagnosis during acute COVID window), the temporal window when measurements were made relative to acute COVID infection, (e.g. pre-COVID window), and by specific clinical domains (e.g. respiratory pathway). At the individual level, we assessed the importance of each variable (indexed across each of the four temporal windows) in predicting PASC. At the temporal level, we assessed the relative importance of each of the four temporal windows (baseline, pre-COVID, acute COVID, and post-COVID) in predicting PASC status. At the level of the clinical domain, we grouped variables based on the following hypothesized mechanistic pathways of PASC: 1) Baseline demographics and anthropometry, 2) Medical visitation and procedures, 3) Respiratory system, 4) Antimicrobials and infectious disease, 5) Cardiovascular system, 6) Female hormones and pregnancy, 7) Mental health and wellbeing, 8) Pain, skin sensitivity, and headaches, 9) Digestive system, 10) Inflammation, autoimmune, and autoantibodies, 11) Renal function, liver function, and diabetes, 12) Nutrition, 13) COVID Positivity, 14) Uncategorized disease, nervous system, injury, mobility, and age-related factors (14). For temporal and clinical domain groupings, we assessed the mean Shapley value of the 10 most predictive features in each group. A full list of our included covariates along with their grouping by temporality and clinical domain is included in our metatable (Supplemental Table 1. Metadata).

## RESULTS

The dataset included 57,672 patients with 9,031 cases, 46,226 controls, and 2,415 patients excluded due to having a PASC diagnosis within 4 weeks of an acute COVID diagnosis. This yielded a final analytic sample of 55,257 participants.

### Predictive performance

Our models accurately predicted PASC diagnosis status among participants in the training sample, with an AUC of 0.947 on a holdout test set (10% of full data).

### Variable importance

#### Individual predictors

We found that the strongest individual predictors (mean absolute Shapley value) of PASC diagnosis were the length of follow-up (0.40), the number of medical visits associated with a diagnosis during the acute COVID period (0.26), data site 10 (0.25), viral lower respiratory infection during the acute COVID period (0.11), age (0.06), total number of medical visits associated with a diagnosis during the baseline period (0.05), total number of medical visits associated with a diagnosis during the post-COVID period (0.05), data site 31 (0.05), never being a smoker (0.04), month of COVID-19 infection (0.03), hospitalization (0.03), data site 11 (0.03), difficulty breathing during the acute COVID period (0.03), data site 39 (0.03), data site 22 (0.03), systemic corticosteroid use during the pre-COVID period (0.02), data site 67 (0.02), missing state information (0.02), and acute respiratory disease during the acute COVID period (0.02) (Figures 1 and 2).

**Figure 1.**
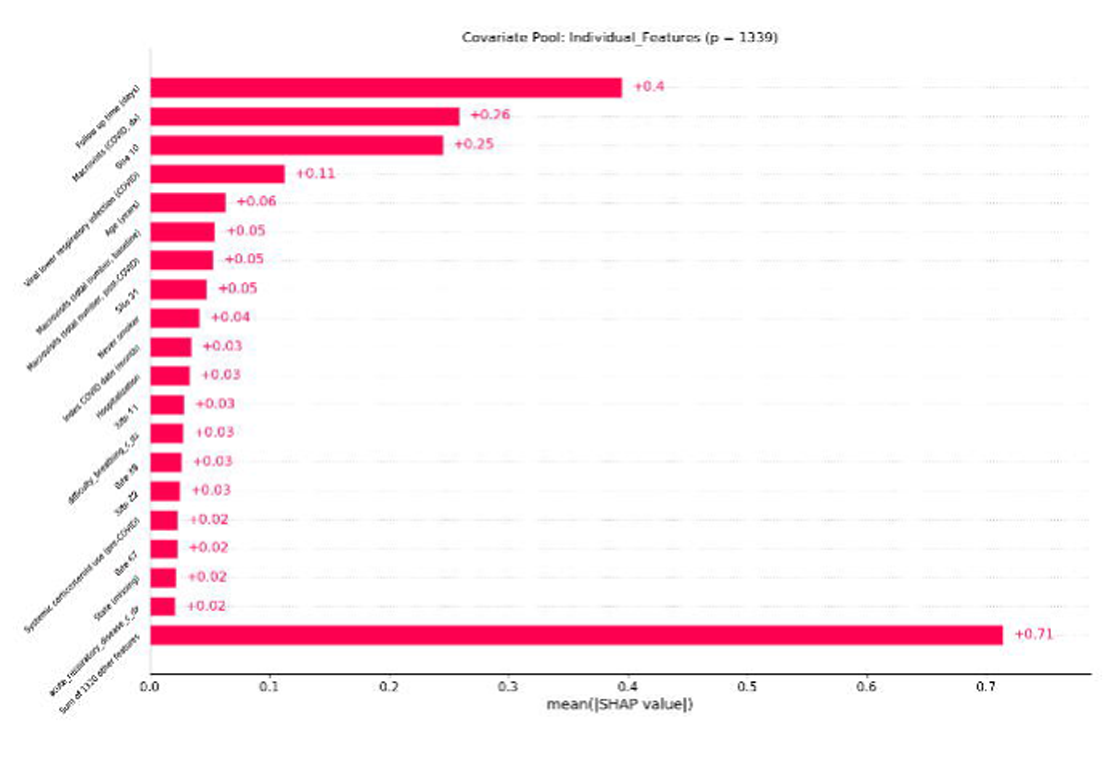
Bar plot of most important model features associated with PASC. For additional information regarding covariates, see metatable.

**Figure 2.**
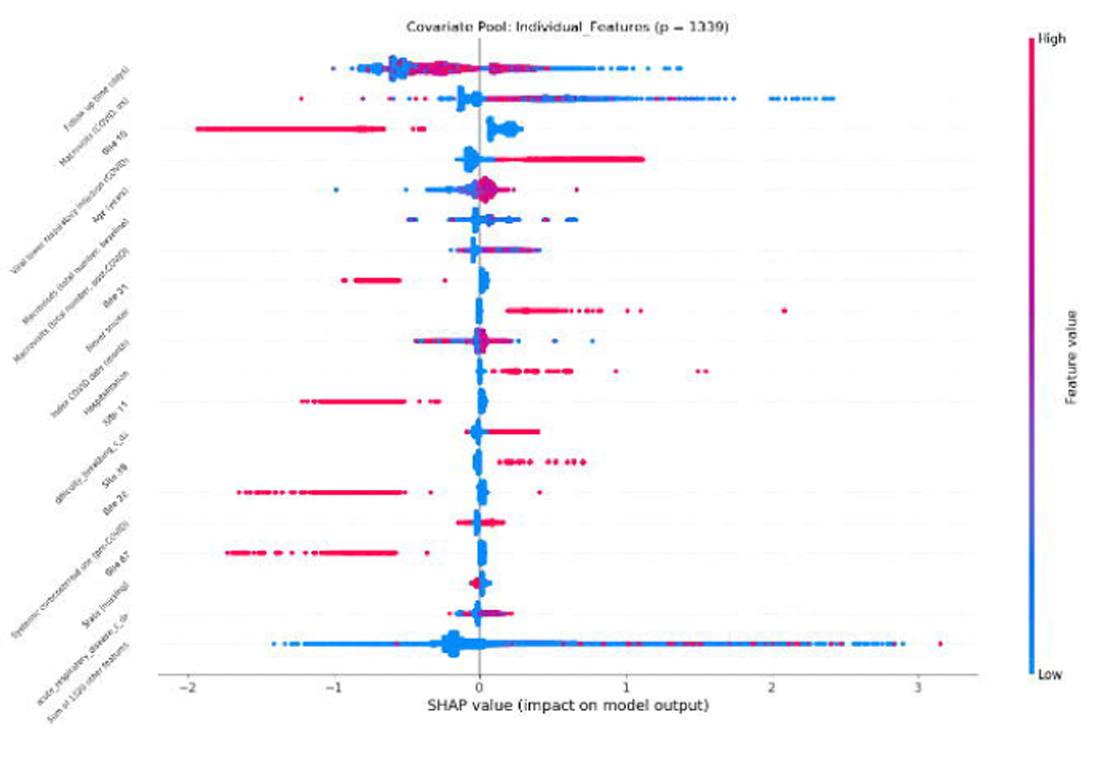
Beeswarm plot of most important model features associated with PASC. For additional information regarding covariates, see metatable.

#### Temporal windows

Baseline and time-invariant characteristics were the strongest predictors of PASC (mean 0.093), followed by characteristics during the acute COVID period (mean 0.049), the post-COVID period (mean 0.013), and the pre-COVID period (0.0098) (Figure 3).

**Figure 3.**
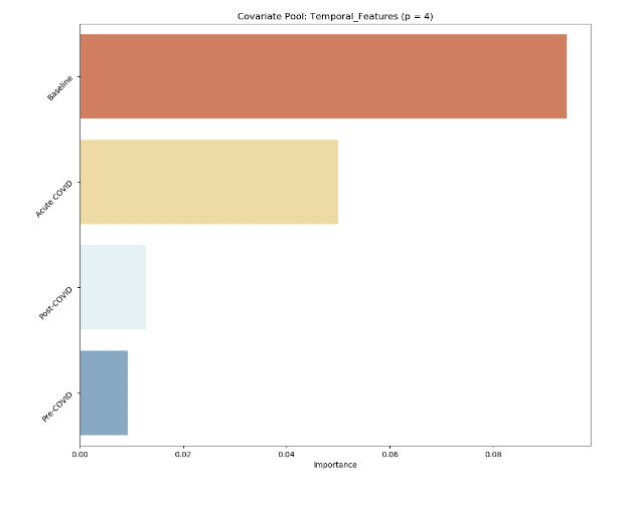
Variable importance by the temporal window. Ranked by the mean absolute Shapley value of the top 10 features in each category. Baseline (prior to t − 37); pre-COVID (t − 37 to t − 7), acute COVID (t − 7 to t + 14), and post-COVID (t + 14 to t + 28), with t being the index COVID date.

#### Clinical domain

We found that medical visitation and procedures included the strongest predictors (mean 0.09), followed by demographics and anthropometry (mean 0.054), respiratory factors (mean 0.023), COVID markers (mean 0.0064), markers of pain (mean 0.0047), cardiovascular factors (mean 0.0039), mental health factors (mean 0.003645), inflammation markers (mean 0.0027), renal and liver factors (mean 0.0026), markers of nutrition (mean 0.0013), factors related to female health and hormones (mean 0.00078), markers of general health and aging (mean 0.00057), infection (mean 0.00027), and digestive (mean 0.00026) (Figure 4).

**Figure 4.**
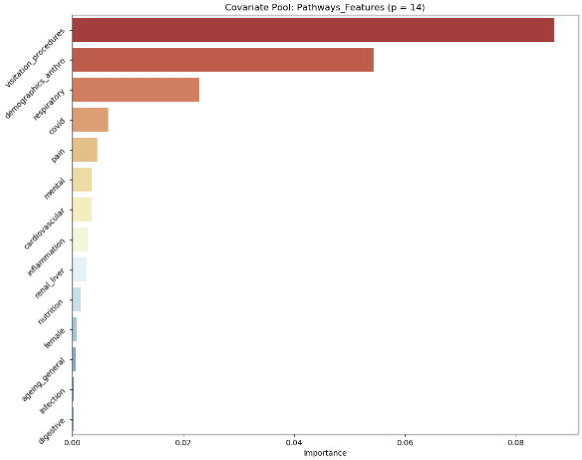
Variable importance by biological pathway. Ranked by the mean absolute Shapely value of the top 10 features (ranked by the same metric) in each category. For additional information regarding covariates, see metatable.

## DISCUSSION

These results provide strong support for 1) the choice of an ensemble learning approach, 2) the specific learners used, 3) how the missing data were handled, and 4) the choice of optimization criteria (maximizing the AUC). Using a similar sample of N3C patients, this Super Learner analysis approach exceeded the AUC reached by Pfaff et al. in predicting PASC diagnosis (AUC 0.95 vs. AUC 0.92, respectively), which used a gradient-boosting prediction algorithm (3). These components are further supported by this model being awarded third place in the NIH Long COVID Computational Challenge (17).

### Individual predictors

We found that the individual predictors most associated with PASC diagnosis were related to medical utilization rate and site of care, such as length of follow-up and data provider ID. These factors may not be causal drivers of PASC incidence and may, rather, indicate incident diagnosis of PASC being more common among those already utilizing medical care, which is consistent with the findings of Pfaff et al. (3). On the other hand, we found that lower tract viral respiratory infection during acute COVID was highly predictive of PASC diagnosis. Previous studies have also linked lower respiratory infection during COVID-19 infection with negative outcomes. A 2022 study found that COVID-19 patients with lower respiratory symptoms experienced worse health outcomes, including supplemental oxygen, mechanical ventilation, and death, compared to patients with upper respiratory symptoms or no respiratory symptoms (18). Lower respiratory infection during acute COVID-19 may be a causal pathway by which acute COVID leads to PASC, although future studies should apply a causal inference framework to evaluate this hypothesis.

### Temporal windows

We found that factors assessed during the baseline period (more than 37 days before COVID-19 diagnosis) were the strongest predictors of PASC diagnosis compared with factors immediately before, during, or after acute COVID-19 infection. This suggests that clinicians may be able to effectively identify who is at risk for PASC based on baseline characteristics, such as preexisting conditions and sociodemographic information. Efforts to develop risk profiles based on these factors should be anchored within a social determinants of health approach, in order to reduce health inequity rather than reinforce systemic inequality (19). Although it should be noted that baseline characteristics included the greatest interval of time and included time-variant factors, such as race. Future analyses should expand on this finding to evaluate the feasibility of predicting individual PASC incidence, rather than diagnosis (which may be subject to bias), using baseline characteristics alone. Additional information regarding this relationship could identify patients at risk for PASC prior to acute COVID-19 and could inform early interventions to prevent PASC.

### Clinical domain

These results are consistent with published literature and highlight the importance of respiratory features (e.g., pre-existing asthma) as important factors in predicting who may develop PASC (2,3). Respiratory factors that may influence individual susceptibility to COVID-19 appear to be important features of acute COVID-19 severity and are key symptoms of PASC (2,3,20). Therefore, future studies should seek to parse the contributions of respiratory symptoms to PASC through the pathways of baseline susceptibility to COVID-19 versus phenotyping of severe COVID-19 in order to improve our understanding of respiratory features as a risk factor for PASC. Despite the range of PASC phenotypes, these findings are consistent with respiratory symptoms (e.g. dyspnea, cough) being the most commonly reported PASC symptoms (14,20). Other clinical domains, such as cardiovascular factors, have similar roles as both markers of susceptibility and severity of COVID-19 and should also be explored further in future studies.

## LIMITATIONS

Our goal for this analysis was to maximize predictive accuracy, rather than to make causal inferences regarding exposure-outcome relationships, therefore we included all predictors prior to four weeks post-COVID (censored window). First, the inclusion of pre-COVID, acute COVID, and post-COVID factors complicate inference regarding whether predictive features (e.g., respiratory factors) reflect vulnerability to acute COVID, COVID symptoms, or early PASC symptoms. Second, this analytic sample was matched 1:4 (PASC:non-PASC), with matching based on pre-COVID medical visitation rate, and this matched sample was drawn from N3C, which is a matched sample of COVID patients and healthy controls. Therefore, this sample may not be representative of a broader population. We note that, for future use of these data, if the prevalence of PASC in the target population is known, and the matching identifier is available, there are methods to calibrate the results to the actual population. Given that was not the case, one might generate results that need to be re-calibrated to the target population of interest. Third, we found measures of medical visitation to be strong predictors of PASC diagnosis. It is plausible that medical visitation may be associated with increased diagnoses of various medical conditions in general, rather than true PASC incidence. However, increased medical visitation may also be an effect of early PASC symptoms. Finally, as is common with electronic health record data, N3C data are heterogeneous with respect to certain outcomes, including biomarker data and PASC diagnosis. A Super Learner-based approach seeks to account for this heterogeneity by modeling underlying patterns of missingness, but residual bias and confounding remain plausible. Overall, this approach enables investigators to make accurate predictions with minimal assumptions despite these data limitations. In order to improve upon the interpretation and clinical applications of these findings, future studies should apply a causal inference approach to evaluate the potential causal impact of individual predictors on the risk of PASC.

## CONCLUSION

These findings provide support for the use of an AUC-maximizing Super Learner approach to predict Long COVID status using N3C data, which may have utility across other binary outcomes in EHR data. These findings highlight the importance of respiratory symptoms, healthcare utilization, and age in predicting PASC incidence. Although further investigation is needed, our findings could support the referral of COVID-19 patients with severe respiratory symptoms for subsequent PASC monitoring. In future work, we plan to investigate predictive performance when only baseline information is used as input to classify PASC, as this provides a practical implementation based on readily available clinical features that could identify participants at risk of PASC prior to COVID diagnosis.

## Supporting information

Supplemental Table 1. Metadata

Appendix 1. Competition Write-Up

## Data Availability

All data analyzed and produced in the manuscript are accessible via the National COVID Cohort Collaborative Data Enclave. A version of the manuscript analysis, using synthetic data rather than de-identified data, can be accessed via GitHub.

https://covid.cd2h.org

https://github.com/BerkeleyBiostats/l3c_ctml/tree/v1

## N3C Attribution

The analyses described in this manuscript were conducted with data or tools accessed through the NCATS N3C Data Enclave https://covid.cd2h.org and N3C Attribution & Publication Policy v 1.2-2020-08-25b supported by NCATS U24 TR002306, Axle Informatics Subcontract: NCATS-P00438-B, and the Bill & Melinda Gates Foundation: OPP1165144. This research was possible because of the patients whose information is included within the data and the organizations (https://ncats.nih.gov/n3c/resources/data-contribution/data-transfer-agreement-signatories) and scientists who have contributed to the on-going development of this community resource [https://doi.org/10.1093/jamia/ocaa196].

## Disclaimer

The N3C Publication committee confirmed that this manuscript (MSID:1495.891) is in accordance with N3C data use and attribution policies; however, this content is solely the responsibility of the authors and does not necessarily represent the official views of the National Institutes of Health or the N3C program.

## IRB

The N3C data transfer to NCATS is performed under a Johns Hopkins University Reliance Protocol # IRB00249128 or individual site agreements with NIH. The N3C Data Enclave is managed under the authority of the NIH; information can be found at https://ncats.nih.gov/n3c/resources.

## Individual Acknowledgements For Core Contributors

We gratefully acknowledge the following core contributors to N3C:

Adam B. Wilcox, Adam M. Lee, Alexis Graves, Alfred (Jerrod) Anzalone, Amin Manna, Amit Saha, Amy Olex, Andrea Zhou, Andrew E. Williams, Andrew Southerland, Andrew T. Girvin, Anita Walden, Anjali A. Sharathkumar, Benjamin Amor, Benjamin Bates, Brian Hendricks, Brijesh Patel, Caleb Alexander, Carolyn Bramante, Cavin Ward-Caviness, Charisse Madlock-Brown, Christine Suver, Christopher Chute, Christopher Dillon, Chunlei Wu, Clare Schmitt, Cliff Takemoto, Dan Housman, Davera Gabriel, David A. Eichmann, Diego Mazzotti, Don Brown, Eilis Boudreau, Elaine Hill, Elizabeth Zampino, Emily Carlson Marti, Emily R. Pfaff, Evan French, Farrukh M Koraishy, Federico Mariona, Fred Prior, George Sokos, Greg Martin, Harold Lehmann, Heidi Spratt, Hemalkumar Mehta, Hongfang Liu, Hythem Sidky, J.W. Awori Hayanga, Jami Pincavitch, Jaylyn Clark, Jeremy Richard Harper, Jessica Islam, Jin Ge, Joel Gagnier, Joel H. Saltz, Joel Saltz, Johanna Loomba, John Buse, Jomol Mathew, Joni L. Rutter, Julie A. McMurry, Justin Guinney, Justin Starren, Karen Crowley, Katie Rebecca Bradwell, Kellie M. Walters, Ken Wilkins, Kenneth R. Gersing, Kenrick Dwain Cato, Kimberly Murray, Kristin Kostka, Lavance Northington, Lee Allan Pyles, Leonie Misquitta, Lesley Cottrell, Lili Portilla, Mariam Deacy, Mark M. Bissell, Marshall Clark, Mary Emmett, Mary Morrison Saltz, Matvey B. Palchuk, Melissa A. Haendel, Meredith Adams, Meredith Temple-O’Connor, Michael G. Kurilla, Michele Morris, Nabeel Qureshi, Nasia Safdar, Nicole Garbarini, Noha Sharafeldin, Ofer Sadan, Patricia A. Francis, Penny Wung Burgoon, Peter Robinson, Philip R.O. Payne, Rafael Fuentes, Randeep Jawa, Rebecca Erwin-Cohen, Rena Patel, Richard A. Moffitt, Richard L. Zhu, Rishi Kamaleswaran, Robert Hurley, Robert T. Miller, Saiju Pyarajan, Sam G. Michael, Samuel Bozzette, Sandeep Mallipattu, Satyanarayana Vedula, Scott Chapman, Shawn T. O’Neil, Soko Setoguchi, Stephanie S. Hong, Steve Johnson, Tellen D. Bennett, Tiffany Callahan, Umit Topaloglu, Usman Sheikh, Valery Gordon, Vignesh Subbian, Warren A. Kibbe, Wenndy Hernandez, Will Beasley, Will Cooper, William Hillegass, Xiaohan Tanner Zhang. Details of contributions available at covid.cd2h.org/core-contributors

## Data Partners with Released Data

The following institutions whose data is released or pending:

### Available

Advocate Health Care Network — UL1TR002389: The Institute for Translational Medicine (ITM) • Boston University Medical Campus — UL1TR001430: Boston University Clinical and Translational Science Institute • Brown University — U54GM115677: Advance Clinical Translational Research (Advance-CTR) • Carilion Clinic — UL1TR003015: iTHRIV Integrated Translational health Research Institute of Virginia • Charleston Area Medical Center U54GM104942: West Virginia Clinical and Translational Science Institute (WVCTSI) • Children’s Hospital Colorado — UL1TR002535: Colorado Clinical and Translational Sciences Institute • Columbia University Irving Medical Center — UL1TR001873: Irving Institute for Clinical and Translational Research • Duke University — UL1TR002553: Duke Clinical and Translational Science Institute • George Washington Children’s Research Institute — UL1TR001876: Clinical and Translational Science Institute at Children’s National (CTSA-CN) • George Washington University — UL1TR001876: Clinical and Translational Science Institute at Children’s National (CTSA-CN) • Indiana University School of Medicine — UL1TR002529: Indiana Clinical and Translational Science Institute • Johns Hopkins University — UL1TR003098: Johns Hopkins Institute for Clinical and Translational Research • Loyola Medicine — Loyola University Medical Center • Loyola University Medical Center — UL1TR002389: The Institute for Translational Medicine (ITM) • Maine Medical Center — U54GM115516: Northern New England Clinical & Translational Research (NNE-CTR) Network • Massachusetts General Brigham — UL1TR002541: Harvard Catalyst • Mayo Clinic Rochester UL1TR002377: Mayo Clinic Center for Clinical and Translational Science (CCaTS) • Medical University of South Carolina — UL1TR001450: South Carolina Clinical & Translational Research Institute (SCTR) • Montefiore Medical Center — UL1TR002556: Institute for Clinical and Translational Research at Einstein and Montefiore • Nemours — U54GM104941: Delaware CTR ACCEL Program • NorthShore University HealthSystem — UL1TR002389: The Institute for Translational Medicine (ITM) • Northwestern University at Chicago — UL1TR001422: Northwestern University Clinical and Translational Science Institute (NUCATS) • OCHIN — INV-018455: Bill and Melinda Gates Foundation grant to Sage Bionetworks • Oregon Health & Science University — UL1TR002369: Oregon Clinical and Translational Research Institute • Penn State Health Milton S. Hershey Medical Center — UL1TR002014: Penn State Clinical and Translational Science Institute • Rush University Medical Center — UL1TR002389: The Institute for Translational Medicine (ITM) • Rutgers, The State University of New Jersey — UL1TR003017: New Jersey Alliance for Clinical and Translational Science • Stony Brook University — U24TR002306 • The Ohio State University — UL1TR002733: Center for Clinical and Translational Science • The State University of New York at Buffalo — UL1TR001412: Clinical and Translational Science Institute • The University of Chicago — UL1TR002389: The Institute for Translational Medicine (ITM) • The University of Iowa — UL1TR002537: Institute for Clinical and Translational Science • The University of Miami Leonard M. Miller School of Medicine — UL1TR002736: University of Miami Clinical and Translational Science Institute • The University of Michigan at Ann Arbor — UL1TR002240: Michigan Institute for Clinical and Health Research • The University of Texas Health Science Center at Houston — UL1TR003167: Center for Clinical and Translational Sciences (CCTS) • The University of Texas Medical Branch at Galveston — UL1TR001439: The Institute for Translational Sciences • The University of Utah — UL1TR002538: Uhealth Center for Clinical and Translational Science • Tufts Medical Center — UL1TR002544: Tufts Clinical and Translational Science Institute • Tulane University — UL1TR003096: Center for Clinical and Translational Science • University Medical Center New Orleans — U54GM104940: Louisiana Clinical and Translational Science (LA CaTS) Center • University of Alabama at Birmingham — UL1TR003096: Center for Clinical and Translational Science • University of Arkansas for Medical Sciences — UL1TR003107: UAMS Translational Research Institute • University of Cincinnati — UL1TR001425: Center for Clinical and Translational Science and Training • University of Colorado Denver, Anschutz Medical Campus — UL1TR002535: Colorado Clinical and Translational Sciences Institute • University of Illinois at Chicago — UL1TR002003: UIC Center for Clinical and Translational Science • University of Kansas Medical Center — UL1TR002366: Frontiers: University of Kansas Clinical and Translational Science Institute • University of Kentucky — UL1TR001998: UK Center for Clinical and Translational Science • University of Massachusetts Medical School Worcester — UL1TR001453: The UMass Center for Clinical and Translational Science (UMCCTS) • University of Minnesota — UL1TR002494: Clinical and Translational Science Institute • University of Mississippi Medical Center — U54GM115428: Mississippi Center for Clinical and Translational Research (CCTR) • University of Nebraska Medical Center — U54GM115458: Great Plains IDeA-Clinical & Translational Research • University of North Carolina at Chapel Hill — UL1TR002489: North Carolina Translational and Clinical Science Institute • University of Oklahoma Health Sciences Center — U54GM104938: Oklahoma Clinical and Translational Science Institute (OCTSI) • University of Rochester — UL1TR002001: UR Clinical & Translational Science Institute • University of Southern California — UL1TR001855: The Southern California Clinical and Translational Science Institute (SC CTSI) • University of Vermont — U54GM115516: Northern New England Clinical & Translational Research (NNE-CTR) Network • University of Virginia — UL1TR003015: iTHRIV Integrated Translational health Research Institute of Virginia • University of Washington — UL1TR002319: Institute of Translational Health Sciences • University of Wisconsin-Madison — UL1TR002373: UW Institute for Clinical and Translational Research • Vanderbilt University Medical Center — UL1TR002243: Vanderbilt Institute for Clinical and Translational Research • Virginia Commonwealth University — UL1TR002649: C. Kenneth and Dianne Wright Center for Clinical and Translational Research • Wake Forest University Health Sciences — UL1TR001420: Wake Forest Clinical and Translational Science Institute • Washington University in St. Louis — UL1TR002345: Institute of Clinical and Translational Sciences • Weill Medical College of Cornell University — UL1TR002384: Weill Cornell Medicine Clinical and Translational Science Center • West Virginia University — U54GM104942: West Virginia Clinical and Translational Science Institute (WVCTSI)

### Submitted

Icahn School of Medicine at Mount Sinai — UL1TR001433: ConduITS Institute for Translational Sciences • The University of Texas Health Science Center at Tyler — UL1TR003167: Center for Clinical and Translational Sciences (CCTS) • University of California, Davis — UL1TR001860: UCDavis Health Clinical and Translational Science Center • University of California, Irvine — UL1TR001414: The UC Irvine Institute for Clinical and Translational Science (ICTS) • University of California, Los Angeles — UL1TR001881: UCLA Clinical Translational Science Institute • University of California, San Diego — UL1TR001442: Altman Clinical and Translational Research Institute • University of California, San Francisco — UL1TR001872: UCSF Clinical and Translational Science Institute

### Pending

Arkansas Children’s Hospital — UL1TR003107: UAMS Translational Research Institute • Baylor College of Medicine — None (Voluntary) • Children’s Hospital of Philadelphia UL1TR001878: Institute for Translational Medicine and Therapeutics • Cincinnati Children’s Hospital Medical Center — UL1TR001425: Center for Clinical and Translational Science and Training • Emory University — UL1TR002378: Georgia Clinical and Translational Science Alliance • HonorHealth — None (Voluntary) • Loyola University Chicago — UL1TR002389: The Institute for Translational Medicine (ITM) • Medical College of Wisconsin — UL1TR001436: Clinical and Translational Science Institute of Southeast Wisconsin • MedStar Health Research Institute — UL1TR001409: The Georgetown-Howard Universities Center for Clinical and Translational Science (GHUCCTS) • MetroHealth — None (Voluntary) • Montana State University — U54GM115371: American Indian/Alaska Native CTR • NYU Langone Medical Center — UL1TR001445: Langone Health’s Clinical and Translational Science Institute • Ochsner Medical Center — U54GM104940: Louisiana Clinical and Translational Science (LA CaTS) Center • Regenstrief Institute — UL1TR002529: Indiana Clinical and Translational Science Institute • Sanford Research — None (Voluntary) • Stanford University — UL1TR003142: Spectrum: The Stanford Center for Clinical and Translational Research and Education • The Rockefeller University — UL1TR001866: Center for Clinical and Translational Science • The Scripps Research Institute — UL1TR002550: Scripps Research Translational Institute • University of Florida — UL1TR001427: UF Clinical and Translational Science Institute • University of New Mexico Health Sciences Center — UL1TR001449: University of New Mexico Clinical and Translational Science Center • University of Texas Health Science Center at San Antonio — UL1TR002645: Institute for Integration of Medicine and Science • Yale New Haven Hospital — UL1TR001863: Yale Center for Clinical Investigation

## Authors statement

Authorship was determined using ICMJE recommendations.

ZB: Generated list of included covariates, drafted writeup, managed competition timeline, attended weekly office hours, and coordinated analysis.

YJ and SS: Screened covariates for inclusion, processed datasets, and developed analysis tools.

HL and JC: Developed analysis workflow for the Enclave, implemented analysis, tuned learners, and designed variable importance framework.

AM, RVP, JC, ML, AH, RCP, and RP: Provided oversight on analysis workflow, gave feedback on drafts and proposed plans, and supported subject matter interpretations.

## Supplemental Materials

Supplemental Table 1. Metadata

Appendix 1. Competition Writeup

