## Appendix 1. Competition Write-Up for "Predicting Long COVID in the National COVID Cohort Collaborative Using Super Learner"

### Predicting PASC in a Matched Sample of Patients with COVID-19

Do you want to make your submission code public as part of the challenge archive? **Yes**

#### Summary Sentence

We predicted individual PASC status given covariate information using Super Learner (an ensemble machine learning algorithm) to learn the optimal, AUC-maximizing combination of gradient boosting and random forest algorithms [1,2]. The filepath of our predictions is “*Submission/workbook-output/STEP2_train_and_predict/Targeted ml team predictions*”.

#### Background/Introduction

As the mortality rate associated with acute COVID-19 incidence wanes, investigators have shifted focus to determining its longer-term, chronic impacts. Post-acute Sequelae of COVID-19 (PASC) is a loosely categorized consequence of acute infection that is related to dysfunction across multiple biological systems [3]. Electronic health record databases, such as the National COVID Cohorts Collaborative (N3C), provide an important tool for predicting, evaluating, and understanding PASC [4].

Given the broad range of factors associated with PASC and the high dimensionality of the N3C Enclave data, modeling methods must be highly flexible. Super Learner (SL) is a flexible, ensemble machine learning algorithm (also known as stacking) that uses cross-validation to learn the optimally weighted combination of a specified set of algorithms [1,2]. The SL is grounded in statistical optimality theory that guarantees for large sample sizes it will perform as well as or better than any algorithm included in the library. Thus, a rich library of learners, with a sufficient sample size, can be difficult to impossible to improve upon under the conditions of the theory. This robustness is supported by numerous applications [2]. The SL can estimate any parameter that can be defined as the optimizer of a particular performance metric (e.g., mean squared error).

Here, we used the SL to estimate the function for predicting PASC diagnosis in COVID-infected patients, and this SL was specified such that it learned the combination of algorithms, including variations of gradient boosting and random forest, that maximized the area under the receiver operator characteristic curve (AUC) [5]. Our set of features for predicting PASC included those previously described in the literature [4], and additional features related to subject-matter knowledge and patterns of missingness. We also investigated the importance of features for predicting PASC across multiple levels, including assessing the importance of each individual feature, and groups of features based on temporality (baseline, pre-COVID, acute COVID, and post-COVID features) and hypothesized biological pathways of PASC.

#### Methods

*Overview and general approach*

We extracted 304 features from N3C data. After indexing across four time periods and transforming features into formats amenable to machine learning analysis, our sample included 1,339 features (see Table 1. Metatable). For continuous features, we included the minimum, maximum, and mean values for each measurement in each temporal window. For binary features, we either included an indicator (when repetition was not relevant) or a count (when repetition was relevant) over each time period and we re-coded categorical variables as indicators. The primary outcome was PASC diagnosis (binary), and we excluded participants who were diagnosed prior to four weeks.

*Temporal windows*

We divided each participant's records into four temporal windows: baseline, which consisted of all records occurring minimum 37 days before the COVID index date ($t$ - 37, where $t$ represents the index COVID date) and all time-invariant factors (such as sex, ethnicity, etc.); pre-COVID, observations falling 37 days prior to 7 days prior to the index date ($t$ - 37 to $t$ - 7); acute COVID, observations falling 7 days prior to 14 days after to the index date ($t$ - 7 to $t$ + 14); and post-COVID, records from 14 to 28 days after the index date ($t$ + 14 to $t$ + 28).

*Features described in the literature*

Earlier this year, Pfaff et al. used gradient-boosting machine learning models (XGBoost) to identify patients at risk for PASC using N3C data [4]. We extracted and transformed key features that were identified by Pfaff et al. These features included 199 previously described factors related to medical history, diagnoses, demographics, and comorbidities [4].

*Temporality*

To account for differences in follow-up, we included as an additional factor a continuous variable for follow-up time, defined as the number of days between the index COVID date and the most recent observation. To account for temporal trends of COVID (such as seasonality and dominant variant), we included categorical (ordinal) covariates for the season and months since the first observed COVID index date.

*Additional features: laboratory measurements, observations, and devices*

We incorporated the laboratory measurements related to anthropometry, nutrition, COVID positivity, inflammation, tissue damage due to viral infection, auto-antibodies and immunity, cardiovascular health, and microvascular disease, which are potential predictors of PASC [6]. We also extracted information about smoking status, alcohol use, marital status, and use of insulin or anticoagulant from the observation table as baseline characteristics of individuals, and we included the number of times a person has been exposed to respiratory devices in each of the four windows from the device table.

*Missing data*

We applied an approach that can be used to predict future observations with missing data, and we did so by creating indicator basis functions that indicate whether, for each variable, the observation was missing (yes/no). By including these (along with filling each missing variable with a 0), we allow the machine to determine what predictive information can be utilized by the missingness process, without relying on a current imputation model. Thus, this indicator allows the pattern of missingness to be a predictor of PASC.

*External data*

We incorporated additional features from the “Covid patient summary table”, within the workbook “Patient Severity Table”. We extracted covariates related to COVID severity, vaccination history, demographics, medical history, and previous diagnoses from before and during acute COVID infection.

*COVID positivity*

We added several measures of COVID severity and persistent SARS-CoV-2 viral load, which are both associated with PASC incidence [6]. We imported measures of COVID severity from the N3C “COVID Severity Table”. In addition, we included 15 measures of COVID infection from laboratory measurements, which provided insights on persistent SARS-CoV-2 viral load. We assessed the duration of COVID viral positivity separately for each laboratory measure of COVID and each temporal window. For participants who had both a positive and negative value of a given test during a temporal window, we took the midpoint between the last positive test and the first negative test as being the endpoint of their positivity. For individuals who had a positive test but no subsequent negative test within that temporal window, we determined their endpoint to be their final positive test plus three days. We included separate missingness indicators in each temporal window for each test, for a positive value for each test, and for a negative value following a positive value to indicate an imputed positivity endpoint.

*Prediction using ensemble machine learning*

We used the SL, an ensemble machine learning method, to learn the optimally weighted combination of candidate algorithms for maximizing the AUC. We reprogrammed the Super Learner in Python in order to capitalize on the resources available in the N3C Data Enclave used for the competition (e.g., PySpark parallelization). This software will be available for broader use beyond the competition. Given the limited time to finish coding for the competition, we used a relatively small ensemble of four learners (a mix of robust parametric models and ml models): 1) logistic regression; 2) L1 penalized logistic regression (with penalty parameter lambda = 0.01); 3) gradient boosting (with n_estimators = 200, max_depth = 5, learning_rate = 0.1); 4) random forest (max_depth = 5, num_trees=20). The original candidate learner library consisted of a large set of candidate learners with different combinations of hyperparameters (e.g. gradient boosting (with n_estimators = [200, 150, 100, 50], max_depth = [3,5,7], learning_rate = [0.05, 0.1, 0.2]). We dropped learners with poor performance in cross-validation (in terms of low AUC and small weights in meta-learning results) and selected the four learners above as a group of efficient learner candidates. Ideally, one should use a much larger learner library with a more comprehensive set of hyperparameters and treat parameter tuning as an inherent step of Super Learning, but we implemented the method described above due to time constraints.

One important decision for optimizing an algorithm is to decide which metric will be used to evaluate the fit and optimize the weighting of the algorithms in the ensemble. We used a novel approach [5] developed specifically for maximizing the area under the curve (AUC). Specifically, we used an AUC maximizing meta-learner with Powell optimization to learn the convex combination of these four candidate algorithms [5]. The SL was implemented with 10-fold cross-validation.

*Variable importance*

The Super Learner is optimized for prediction but provides no direct parameters that could be considered measures of the importance of each predictor in the context of all other predictors utilized (i.e., no direct measure of variable importance). In this section, for the sake of computational efficiency, we use the discrete SL selector (the single candidate learner with the highest cross-validated AUC, in this case, a gradient-boosting learner) instead of the entire ensemble SL. We first used a general approach (for any machine learning algorithm) known as Shapley values [7]. We generated these values within three groupings of predictors for ease of interpretability: individual features (e.g. cough diagnosis during acute COVID window), temporal window (relative to acute COVID infection, e.g. pre-COVID window) when measurements were made, and by specific biological pathways (e.g. respiratory pathway). At the individual level, we assessed the importance of each variable (indexed across each of four time windows) in predicting PASC. At the temporal level, we assessed the relative importance of each of the four temporal windows (baseline, pre-COVID, acute COVID, and post-COVID) in predicting PASC status. At the level of the biological pathway, we grouped variables based on the following hypothesized mechanistic pathways of PASC: 1) Baseline demographics and anthropometry, 2) Medical visitation and procedures, 3) Respiratory system, 4)  Antimicrobials and infectious disease, 4) Cardiovascular system, 5) Female hormones and pregnancy, 6) Mental health and wellbeing, 7) Pain, skin sensitivity, and headaches, 8) Digestive system, 9) Inflammation, autoimmune, and autoantibodies, 10) Renal function, liver function, and diabetes, 11) Nutrition, 12) COVID Positivity, 13) Uncategorized disease, nervous system, injury, mobility, and age-related factors [6]. A full list of our included covariates along with their grouping by temporality and biological pathway is included in our metatable (Table 1). Our process for analysis and interpretation followed a Targeted Learning strategy [8,9,10].

#### Conclusion/Discussion

*Predictive performance*

Our models accurately predicted PASC diagnosis status among participants in the censored training sample, with an AUC of 0.947 on a holdout test set (10% of full data).  These results provide strong support for 1) the choice of an ensemble learning approach, 2) the specific learners used, 3) how the missing data was handled, and 4) the choice of optimization criteria (maximizing the AUC).

*Variable importance*

We found that the strongest individual predictors (mean absolute Shapley value) of PASC diagnosis were the length of follow-up (0.38), the number of macrovisits associated with a diagnosis during the acute COVID window (0.25), data partner ID 124 (0.24), viral lower respiratory infection during the acute COVID window (0.11), and age (0.07) (Figures 1 and 2).

Regarding the temporal assessment of variable importance, baseline and time-invariant characteristics were the strongest predictors of PASC, followed by characteristics during the acute COVID window (Figure 3). Although it should be noted that baseline characteristics included the greatest interval of time and included some time-variant factors that were not linked to any specific time point. This suggests that clinicians may be able to effectively identify who is at risk for PASC based on baseline characteristics and COVID infection symptoms.

For the biological pathways variable importance analysis, we found that medical visitation and procedures included the strongest predictors, followed by respiratory factors, demographics and anthropometry, measures of liver and renal function, COVID markers, and cardiovascular factors (Figure 4). These results are consistent with published literature and highlight the importance of respiratory features (e.g., asthma) as important factors in predicting who may develop PASC, which is consistent with the fact that SARS-CoV-2 is a respiratory virus [3,4]. Respiratory factors can influence individual susceptibility to COVID-19, are important features of acute COVID-19 severity, and are key symptoms of PASC [3,4,11]. Therefore, future studies should seek to parse the contributions of respiratory symptoms to PASC through the pathways of baseline susceptibility to COVID-19 versus phenotyping of severe COVID-19 in order to improve our understanding of respiratory features as a risk factor for PASC. Despite the range of PASC phenotypes, these findings are consistent with respiratory symptoms (e.g. dyspnea, cough) being the most commonly reported PASC symptoms [4,10]. Other biological pathways, such as cardiovascular factors, have similar roles as both markers of susceptibility and severity of COVID-19 and should also be explored further in future studies.

*Limitations*

Our goal for this analysis was to maximize predictive accuracy, rather than to make causal inferences regarding exposure-outcome relationships, therefore we included all predictors prior to four weeks post-COVID (censored window). The inclusion of pre-COVID, acute COVID, and post-COVID factors complicates inference regarding whether predictive features (e.g., respiratory factors) reflect vulnerability to acute COVID, COVID symptoms, or early PASC symptoms. This analytic sample was matched 1:4 (PASC:non-PASC), with matching based on pre-COVID medical visitation rate, and this matched sample was drawn from N3C, which is a matched sample of COVID patients and healthy controls. Therefore, this sample may not be representative of a broader population. We note for future use of these data, if the prevalence of PASC in the target population is known, and the matching id is available, there are methods to calibrate the results to the actual population. Given that was not the case, one might generate results that need to be re-calibrated to the population of interest.

We found measures of medical visitation to be strong predictors of PASC diagnosis. It is plausible that medical visitation may be associated with increased diagnoses in general, rather than true PASC incidence, although increased medical visitation may be an effect of early PASC symptoms.

*Summary*

These findings highlight the importance of respiratory symptoms, healthcare utilization, and age in predicting PASC incidence, which is consistent with Pfaff et al. [4]. Although further investigation is needed, this supports the referral of COVID-19 patients with severe respiratory symptoms for subsequent PASC monitoring. In future work, we plan to investigate predictive performance when only baseline information is used as input to classify PASC, as this provides a practical implementation based on readily-available clinical features that could identify participants at risk of PASC prior to COVID diagnosis.

### Workbook Specifications

*Environment*:

a) For data processing: profile-high-memory (only required for final merging script, default profile is sufficient for preceding data processing scripts)

b) For model training: profile-high-memory

c) For variable importance: profile-high-memory (custom, with shap module)

*Libraries*:

a) For data processing: SQL, pyspark, pandas

b) For model training: numpy, pandas, sklearn, scipy.optimize, pyspark.ml.feature, pyspark.sql.functions, pyspark.ml.classification, pyspark.sql.types

c) For variable importance: part b) plus: shap, matplotlib, seaborn

*Estimated Runtime:*

a) For data processing: ~30 minutes

b) For model training: ~1.2 hours for model fitting (10-fold cv), ~30 minutes for fitting models on full data and generating predictions for test data.

c) For variable importance: ~10-20 minutes

#### Workbook References

a) For feature preprocessing:

a.1) feature preprocessing on individual raw tables (2_medication, 3_diagnosis, 4_lab_measures, 5_comorbidity, 6_covid_measures, 7_device, 8_observation)

E.g. lab measurements

- *Workbook name: STEP1_feature > 4_lab_measures*
- *Transform name(s)*: measurement_person, pre_pre_measurement, pre_measurement, covid_measurement, post_measurement, four_windows_measure, lab_measures_clean
- *Description*: These transformations take the raw OMOP table measurement and the training cohort to get their measurements of interests in each of the four windows. For each window, minimum, maximum and mean values of those measurements were calculated for each individual.
- *Input Data: measurement, Feature_table_builder*

*a.2) feature engineering to combine the processed tables*

- *Workbook name:* STEP1_feature > 9_combine
- *Transform name(s)*: severity_table, pre_post_med_final_distinct, add_labels, condition_rollup, parent_condition_rollup, final_rollups, add_alt_rollup,

pre_post_dx_final, count_dx_pre_and_post, feature_table_all_patients_dx_drug, metatable
- *Description*: Pivot the individual processed tables and combine their together. Also conducted encoding, null interpolation and other necessary transformations to generate the final dataset
- *Input Data:* *concept, pre_post_dx_count_clean, concept_ancestor, long_covid_patients, Feature_table_builder, pre_post_med_count_clean, Lab_measures_clean, Comorbidity_counts, Covid_measures, Device_count_clean, Obs_person_pivot, Covid_patient_summary_table*

b) For model training:

b.1) model training on subset (90% of ~55k) and evaluation on holdout test set (10%)

- *Workbook name: STEP2_train_and predict*
- *Transform name(s)*: train_dat, test_dat, analytic_train_dat, analytic_test_dat, train_sl_sub, eval_sub_fit
- *Description*: The first four transforms randomly split the full training data into training and holdout test sets. The train_sl_sub fits SL with the training set and outputs a df showing weights and cv AUC of each candidate learner. The eval_sub_fit fits each learner on the training set, combines them with weights from train_sl_sub, and finally evaluates the ensemble SL predictions on the holdout test set.
- *Input Data:* Feature_table_all_patients_dx_drug

b.2) model training on full set (n=~55k) and predicting on real test set (n=300)

- *Workbook name: STEP2_train_and predict*
- *Transform name(s)*: analytic_full_train, analytic_final_test, train_sl, targeted_ml_team_predictions
- *Description*: The first two transforms prepare the full training data and test data respectively. The train_sl fits SL with the full training data and outputs a df showing weights and cv AUC of each candidate learner. The targeted_ml_team_predictions fits each learner on the full training data, combines them with weights from train_sl_sub, and finally uses the ensemble SL to make predictions on the test data for submission.
- *Input Data:* Feature_table_all_patients_dx_drug, feature_table_final_testing_set

c) For variable importance:

- *Workbook name: STEP3_variable_importance*
- *Transform name(s)*: analytic_full_train, vim_shap, plots_formatted
- *Description*: The first transform prepares the full training data. The vim_shap fits a gradient boosting (the best learner in the ensemble SL, also known as the discrete SL) with the full training data and generates Shapley plots. The plots_formatted is the same as the vim_shap, it additionally adds some tentative formats to the plots for this project.
- *Input Data:* Feature_table_all_patients_dx_drug, Metatable


**Authors statement**

ZB: Generated list of included covariates, drafted writeup, managed competition timeline, attended weekly office hours, and coordinated analysis.

YJ and SS: Screened covariates for inclusion, processed datasets, and developed analysis tools.

HL and JC: Developed analysis workflow for the Enclave, implemented analysis, tuned learners, and designed variable importance framework.

AM, RVP, JC, AH, and RP: Provided oversight on analysis workflow, gave feedback on drafts and proposed plans, and supported subject matter interpretations.
